# Socioeconomic Inequities in Early Kidney Care Concentrate at Late Initial Clinical Detection Rather Than Planned Care: A Population-Based Study

**DOI:** 10.64898/2026.10.30.26366923

**Authors:** Eilidh Cowan, Magdalena Rzewuska Diaz, Samira Bell, Simon DS Fraser, Audrey Hughes, Buse Keskindag, Shona Methven, Mintu Nath, Dorothea Nitsch, Nicole Scholes-Robertson, Rachel Smith, Philippa Traill, Tom Blakeman, Simon Sawhney

## Abstract

**Introduction:** Socioeconomic inequities in chronic kidney disease (CKD) outcomes are recognised and greater at early stages, but where exactly inequities arise within early care remains unclear. This distinction is critical: if disparities occur before clinical recognition, strategies focused on management after diagnosis may have limited impact.

**Methods:** We conducted a population-based cohort study linking healthcare and Census data for adults in North-East Scotland, 2011-2020. We examined sequential components of early kidney care from initial clinical detection (eGFR <60 mL/min/1.73 m²). Exposures were household and neighbourhood deprivation and living circumstances (rurality, living alone, mental health). Components of early kidney care were: prior surveillance; presentation (including late/unmonitored); confirmation/monitoring; treatment initiation; and unplanned emergency care. Adjusted risk ratios were estimated using modified Poisson models. Mortality was assessed up to 10 years using Cox regression with separate short-term (<1 year) and long-term estimates.

**Results:** Among 44,252 individuals, inequities were most pronounced at initial clinical detection. Individuals from deprived households were more likely to present late (adjusted RR 1.69, 95% CI 1.48–1.93). Inequities in planned monitoring after presentation were less evident (RR 1.00, 0.95– 1.05). Urine albumin testing was infrequent among those with cardiovascular risk factors. Late presentation was also associated with increased mortality (adjusted HR 1.78, 1.70–1.87), persisting beyond the first year.

**Conclusions:** Socioeconomic inequities in CKD are greatest at or before initial clinical detection, with late presentation a measurable indicator. Emphasis on primary care monitoring and prescribing is unlikely to reduce disparities if early detection and awareness are not also addressed.

**Key messages:** *What is already known on this topic:* Socioeconomic inequities in chronic kidney disease (CKD) outcomes are recognised and greater at early stages, but there is limited evidence of where these inequities arise within the early care pathway. If disparities occur before clinical detection, strategies focused on improving management of already known patients after diagnosis may have limited impact.

*What this study adds:* Deprived populations were substantially more likely to present late, whereas differences in subsequent monitoring and treatment were small. Late presentation was strongly associated with mortality, identifying a high-risk group that reflects upstream inequities in detection and access.

*How this study might affect research, practice or policy:* These findings challenge strategies that focus solely on optimising monitoring and prescribing for patients already known to services. Efforts to improve CKD outcomes should prioritise earlier detection and engagement in underserved communities. Late presentation may serve as a measurable indicator of inequitable access and system performance in early kidney care.

## Introduction

The health and care of people with kidney diseases are inequitable, with socioeconomically deprived communities experiencing worse outcomes^1^. This is especially evident in the earliest or mildest stages of chronic kidney disease (CKD), although the underlying reasons remain unclear ^2, 3^. Early-stage kidneycare is critical because emerging cardiovascular-kidney-metabolic (CKM) treatments rely on timely detection, confirmation, and clinical evaluation^4^. Prompt treatment offers greater potential to modify disease trajectories before complications become unavoidable^5^. However, it remains unclear whether inequities arise primarily before patients become clinically visible, or during subsequent care.

Recent advances in CKD treatments have prompted calls for action “earlier” in the disease pathway, and for a shift in focus towards the community^6, 7^. Innovations include integrated primary-secondary care platforms to optimise triage, prescribing and referrals once people with early CKD become visible within the health system^8^. However, “early” as a phase of care is conceptually complex. In nephrology, the terms *early*/*mild*, and *late*/*severe* are often used interchangeably, but these may not align from an equity perspective^9, 10^. “Early” may refer to a younger age, milder severity, shorter observable disease duration, or timely initial detection when treatments remain effective. We therefore conceptualise early kidney care as the point of initial clinical visibility rather than disease severity alone. At this point, guidelines recommend that initial detection should prompt a sequence of steps to enable preventive care and reduce unplanned emergencies^11–14^. Each step may be differentially affected by socioeconomic disadvantages, yet evidence remains limited.

Most studies of inequities in CKD care have focused on neighbourhood deprivation or later disease stages^2, 15, 16^, including individuals already under nephrology care or populations defined by specific comorbidities^17–24^. Within early care, the crucial timeliness of initial detection occurs has received especially little attention, particularly in relation to multiple (individual, household, neighbourhood) layers of socioeconomic disadvantage. It also remains unclear whether inequity spans across all components of early kidney care (surveillance, detection, confirmation, treatment initiation, and prevention of acute unscheduled care) or is concentrated within specific components of care or populations. Without this understanding, current strategies may overemphasise downstream care while neglecting critical upstream barriers, risking widening disparities where treatment may be most effective^25^.

We therefore systematically evaluated equity across sequential components of early kidney care to identify where along the early CKD pathway inequities emerge, where they are greatest, and which populations are most affected.

## Methods

### Study design

GLOMMS-CORE is a population study of linked kidney health data (including primary and secondary care laboratory tests) and census records to study the health and living circumstances of all adults in the North of Scotland (Grampian)^3^. The dataset contains linked real-world health episode information, medications, laboratory data from all clinical locations, and responses to a mandatory national Census which was completed by 458,897 adult residents in 2011. This analysis used data from the KINDER study (Kidney Inequalities: Needs Data Experiences Response), a Scottish Government Chief Scientist Office-funded mixed-methods study involving a critical realist approach to explore how and why health inequities are experienced (CSO grant reference HIPS 23/17). Elsewhere the data have been used to evaluate the mortality outcomes of people with kidney disease in deprived households and neighbourhoods^3^.

The study idea was conceived by our patient partners including co-author AH (Chair, Grampian Kidney Patient Association) and was co-designed with patient and public contributors. A panel of four patient and public partners met regularly with the research team and contributed to the interpretation of findings and preparation of study outputs. The statistical modelling approach was developed and interpreted through deliberations between qualitative and quantitative analysts, the study leads (primary care, nephrology, a patient lead, and a community-based counsellor), and methodological, clinical, and public and patient partners. This collaborative approach ensured that findings were informed by clinical, methodological, lived-experience, and community perspectives.

Through a review of applicable clinical guidelines and confirmation from clinical experts (kidney, primary care and public health professionals) we identified four components of early kidney care that align with UK relevant indicators and CKD guidelines^11, 12^. We included a fifth component (unplanned care) in recognition of literature that uses unscheduled emergency department attendances and avoidable hospital admissions as a sign of failure of navigation^13,14^. We operationalised and evaluated each component across household and neighbourhood levels of deprivation and disadvantage (table 1 and below). This approach enabled simultaneous evaluation of multiple sequential components of early kidney care across different levels of socioeconomic disadvantage.

1. prior surveillance of high-risk individuals;
2. circumstances and severity at initial presentation;
3. initial confirmation and subsequent monitoring;
4. initiation of treatment; and
5. unplanned care.

**Table 1:** Sequential components of early kidney care.

| Domain of clinical care | Care process | Explanation | n / N | % |
|---|---|---|---|---|
| (1) Prior surveillance of high-risk individuals | Prior outpatient creatinine among high risk | Those newly presenting with kidney impairment who have a prior high-risk feature for kidney disease (diabetes or a vascular disease including coronary disease, heart failure, hypertension, atrial fibrillation, stroke) and have evidence of prior serum creatinine surveillance in the community (within 15 months, i.e. a 12 month annual health check ± 3 months) | 16401 / 18746 | 87.5 |
|  | Prior ACR among high risk | Annual screening for proteinuria as evident (from ACR or PCR tests) in the prior 15 months. | 4720 / 18746 | 25.2 |
| (2) Circumstances and severity at initial presentation | Late presentation eGFR <45 (excluding AKI) | Evidence of a potentially delayed and unmonitored presentation based on a first test of kidney impairment that has skipped CKD stage 3A (i.e. eGFR already <45) in the absence of evidence of AKI or AKD. | 6042 / 39780 | 15.2 |
|  | Inpatient presentation (excluding AKI) | First presentation of kidney impairment in the context of a hospital admission (rather than in the community) in the absence of evidence of AKI or AKD. | 10074 / 39780 | 25.3 |
| (3) Initial confirmation and subsequent monitoring | Repeat test 14d | Evidence of confirmation of low eGFR with a repeat test within 2 weeks among survivors who are not in an inpatient setting. | 9375 / 29823 | 31.4 |
|  | Test for chronicity after 90d | Evidence of testing for sustained chronicity with a repeat blood test after 90 days and within 365 days among survivors after presentation. | 26467 / 38462 | 68.9 |
| (4) Initiation of treatment | Available ACR test | Availability of a previous ACR test from which to determine need for RAASi treatment | 9755 / 38462 | 25.4 |
|  | RAASi among indicated | RAASi use within 365 days where indicated based on most recent ACR status (diabetes & urine ACR >3; or hypertension & urine ACR >30; or urine ACR >70 mg/mmol) | 1057 / 1523 | 69.4 |
| (5) Unplanned care | Emergency department attendance | Presentation in emergency department (ED) within 365 days after presentation among survivors with ED data linkage* (regarded in literature as representing a failure of navigation) | 4131 / 25084 | 16.5 |

\*Domain (5) covered individuals from 2014 and later due to lack of emergency deprtment data availibilty prior to 2014.
Note: These components are based on references <sup>11-14</sup> and in particular the National Institute for Health and Care Excellence guidelines for chronic kidney disease NG203

### Population – new kidney impairment

To mirror the point of an initial presentation and evaluation of impaired kidney function, we studied all adult residents of Grampian (aged 18 and over) with new (incident) evidence of reduced kidney function (eGFR <60 ml/min/1.73m^2^), 2011-2020, based on the race-free version of the 2009 CKD EPI equation^26^. As previously described, we included a two year (2009-2010) initial exclusion “burn-in” phase (i.e. exclusion of individuals who already have evidence of eGFR <60) to avoid mixing of incident and prevalent patients^3^.

### Exposures – deprivation and social circumstances

The main exposure of the study was household deprivation measured using the National Statistics Socio-economic classification (NS-SEC) based on the occupation of the household reference person as recorded in the Census^27^. NS-SEC is the key measure of deprivation in the Census and is central to policy decisions in UK society. For additional exposures, we reported neighbourhood deprivation in quintiles of the Scottish Index of Multiple Deprivation (SIMD)^28^. This was determined from the resident postcode at presentation, and represents an aggregate measure given to a neighbourhood based on seven domains: income, employment, health, access to services, education, housing and crime. Ǫuintile 1 refers to the most deprived 20% of neighbourhoods in Scotland and quintile 5 the most affluent 20%. We additionally examined rurality using postcode-based classification (Scottish Urban Rural Classification), with a pre-specified/*a priori* distinction between rural residents with and without access to a car^29^. We also included self-reported mental health conditions and living status (living alone vs not) as additional exposures.

### Outcomes – presenting circumstances and processes of early kidney care

Details of the five components of early kidney care are summarised in table 1, each mapped to recommended care processes and measures^11–14^. For (1), prior monitoring of annual kidney blood (creatinine, eGFR) and urine (albumin to creatinine ratio, ACR) tests, analyses were restricted to individuals with diabetes or vascular disease, consistent with guideline-defined high-risk groups. Where we refer to outpatient monitoring, this covers those who were not hospital inpatients at the time of test, irrespective of the community setting or practitioner who organised the blood test. For (2), presenting circumstances of initial detection including late presentation, we excluded those with evidence of AKI according to KDIGO creatinine change criteria^30^. For (3), confirmatory testing within 14 days (recommended in guidelines to identify acute illness) we excluded individuals who were already inpatients (and therefore did not have to overcome access barriers) or who had pre-existing evidence of AKI. For (4), prescription of a renin-angiotensin aldosterone system inhibitor (RAASi), analyses were limited to individuals with a clinical indication based on diabetes or hypertension and a corresponding level of albuminuria. Because the clinical indication for RAASi is contingent on prior measurement of albuminuria, inequities may arise at multiple dependent steps (e.g. access to testing of albuminuria, or prescribing of RAASi if albuminuria is elevated). As the study period pre-dated guideline recommendations for sodium–glucose co-transporter 2 (SGLT2) inhibitors and glucagon-like peptide-1 (GLP-1) agonists, these treatments were not included in the analysis. For (5), unplanned care attendance, we restricted to emergency department use over the subsequent year among survivors.

### Outcomes – mortality

For the mortality outcome, people were followed up to ten years from the date of presentation to the date of death, outmigration, or end of available follow-up (2021).

### Covariates

Covariates included age, sex, and comorbidities derived from routinely collected hospital medical records. Comorbidities included in the analyses were prior history of cancer, chronic obstructive pulmonary disease, coronary heart disease, diabetes, heart failure, hypertension, liver disease, peripheral arterial disease, atrial fibrillation, and stroke based on hospital episode ICD-10 codes available back to 2004, as commonly associated with CKD and as described in our previous work^3^.

### Analyses

We described characteristics for all people in the cohort, including age, sex, deprivation, comorbidities and social circumstances. We reported the number of people at risk for each care process measure, and the proportions of people who received care within each exposure group.

For estimating case-mix adjusted comparisons between socioeconomic categories, odds ratios from multivariable logistic regression can be difficult to interpret and may appear to overstate associations when outcomes are common. We therefore estimated case-mix adjusted risk ratios using modified Poisson regression, implemented as generalised linear models with a Poisson distribution and log link function, together with robust (sandwich) variance estimators to obtain valid standard errors for binary outcomes^31^. Case-mix adjusted proportions were estimated using model-based predicted probabilities derived from average marginal effects over the observed covariate distribution. To assess model specification, we also fitted multivariable logistic regression models, which estimated near identical case-mix adjusted proportions. All models were adjusted for age (continuous), sex, and the comorbidities outlined above. As supplementary analyses determined *a priori*, we tested for interactions by working age status (16-65 years vs >65 years), sex, and diabetes, using robust Wald tests based on joint tests of interaction terms in the modified Poisson models.

To further understand the health implications of presenting “late and unmonitored” for health outcomes, we fitted Cox models for mortality from the point of diagnosis comparing three groups who: (1) those first presented with eGFR <60 but ≥45 ml/min/1.73m^2^ (i.e. an early and mild initial kidney presentation); (2) those with a previous eGFR <60 who now newly presented with eGFR <45 ml/min/1.73m^2^ (i.e. severe but monitored from a previous early and mild presentation); and (3) those with no previous evidence of kidney impairment who now presented at the same time with both eGFR <60 and <45 ml/min/1.73m^2^ (i.e. a late and unmonitored presentation). Models were adjusted for age, sex, and comorbidities as listed above. In addition, recognising the clinical continuity between rapid CKD progression and AKI, again we excluded any individuals with AKI based on KDIGO creatinine change criteria. We further assessed for non-proportionality over follow-up by splitting follow-up into short (<1yr) and long terms (1-5 years). Analyses were conducted in R version 4.4.1, and Stata version 19.

## Results

### Overall cohort description

44,252 patients newly presented with kidney impairment (eGFR <60) and were included in evaluation of at least one care process in the study (table 2). Delivery of guideline-recommended care processes was low (table 1). 30% were of working age (≤65 years), and 55% were female. 15% of individuals initially presented late, and 25% as inpatients. 25% of high-risk individuals had a prior urine ACR checked as part of regular surveillance before presentation, and subsequent ACR testing over the following year resulted in similarly low ACR availability for treatment decisions (25%). Among those with established eligibility based on ACR and comorbidity, RAASi prescribing occurred in 69%. Of guideline recommended confirmatory tests, 31% had a blood test within 14 days to exclude acute illness, and 69% had a test to establish chronicity after 90 days. Over the subsequent year, 17% of individuals received unscheduled emergency care.

**Table 2:**
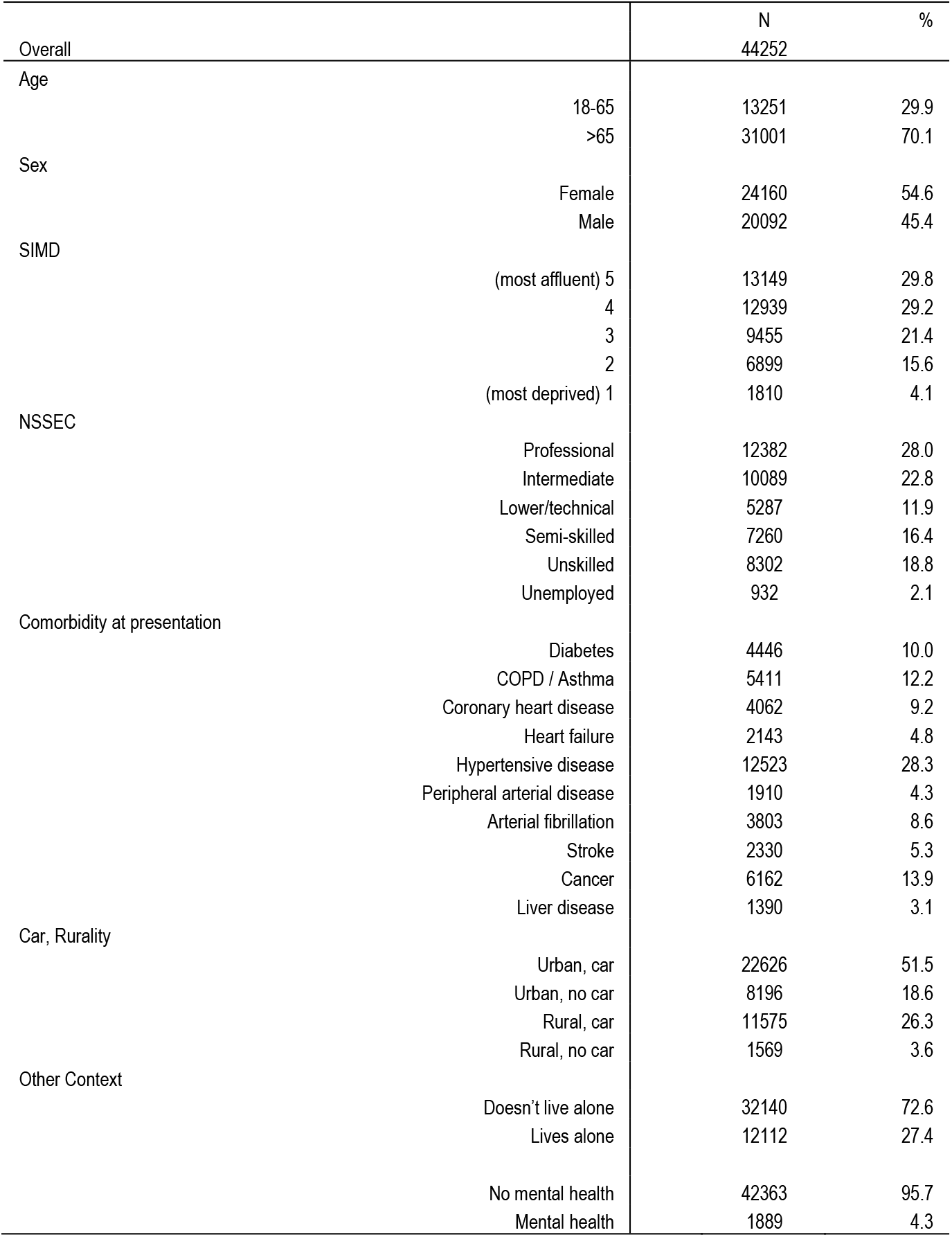
Cohort characteristics.

### Socioeconomic disadvantage and initial presenting circumstances

Multivariable adjusted risks and risk ratios (RR) and confidence intervals (CI) of presenting care processes and circumstances are reported in figure 1 and supplementary table 1. People in deprived communities were less likely to have blood creatinine surveillance for high-risk indications although this difference was modest (RR and 95% CI for most deprived households vs professionals 0.93 [0.89-0.98]). A similar difference was not observable for ACR testing, which occurred substantially less frequently among those with cardiovascular vs diabetes risk factors (10% vs 75%). In contrast, deprivation was associated with markedly increased risks of a late / unmonitored initial presentation (RR deprived vs affluent households 1.69 [1.48-1.93]), or of presenting in inpatient circumstances (RR 1.48 [1.34-1.63]). These patterns of late and inpatient presentations were similar for those who lived alone (respective RRs 1.34 [1.27-1.41], 1.26[1.21-1.30]) and those without access to a car, irrespective of the rurality of their neighbourhood. The patterns were also evident but less marked for those with self-reported mental health conditions (RRs 1.22 [1.10-1.35], 1.21[1.13-1.31]).

**Figure 1:**
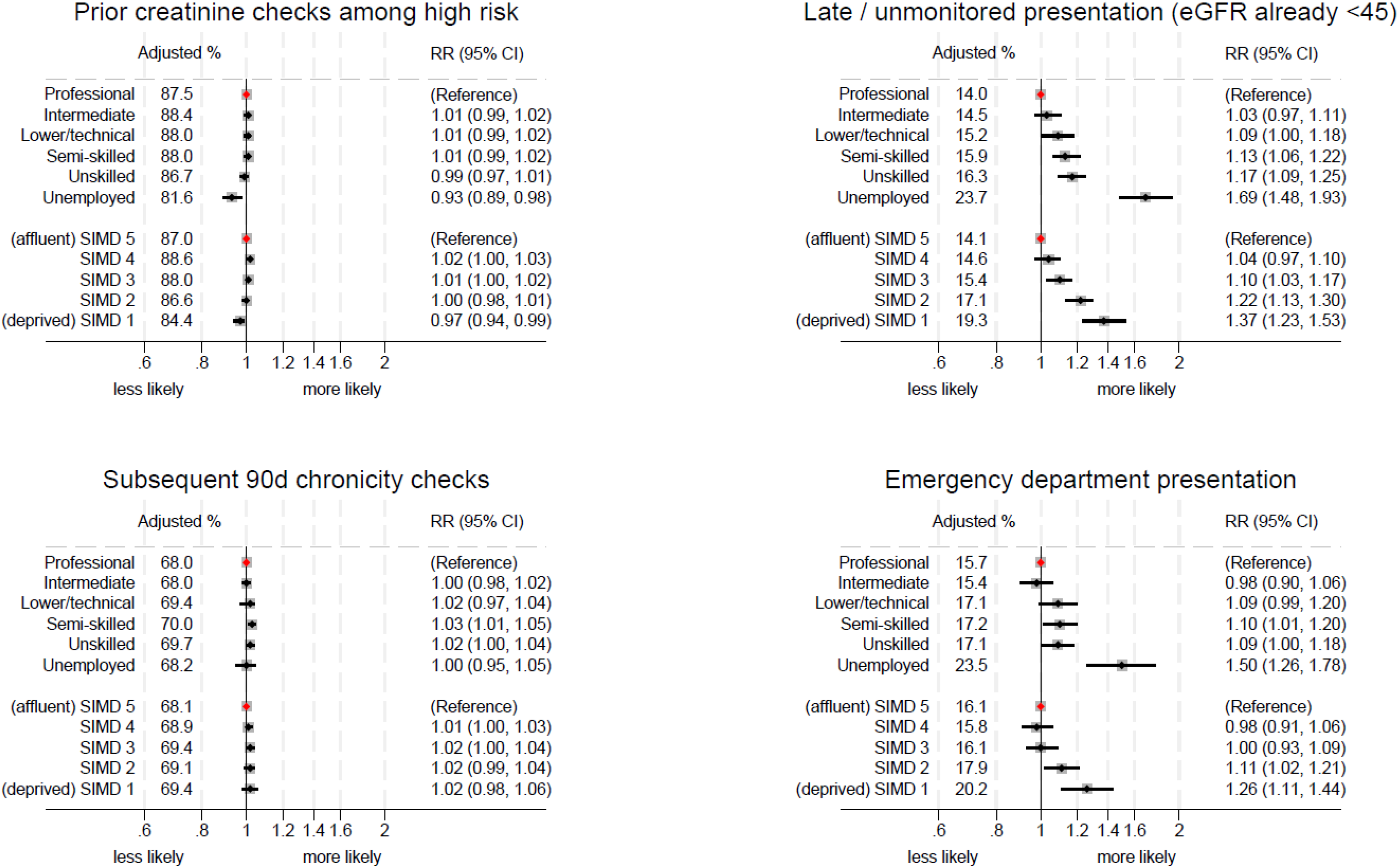
Relationship between processes of care, household socioeconomic classification and deprivation

### Socioeconomic disadvantage and subsequent care

There was no consistent evidence that people in deprived neighbourhoods or households were less likely to receive planned care processes such as 14 and 90-day checks respectively to rule out AKI, and assess chronicity (RRs for household 0.92 [0.80-1.06], 1.00 [0.95-1.05]), nor was there evidence of a substantial difference based on rurality, access to car, living alone or mental health (figure 1 and supplementary table 2). In contrast, there were substantial differences in risk of subsequent unscheduled attendance of the emergency department for deprived households (RR 1.50 [1.26-1.78]) and those who reported mental health conditions (RR 1.54 [1.38-1.72]). Overall, figure 1 demonstrates an evident graded relationship between increasing deprivation and late presentation and emergency department use, but less evidence of difference in surveillance and chronicity monitoring at household or neighbourhood levels.

In supplementary analyses, there was not evidence that the pattern of these overall associations was different for those with vs without diabetes or males vs females (p values for interaction terms all >0.05). There was statistical evidence of interaction with working vs older age (some p values <0.05) and stratified analyses were undertaken. While some findings were more pronounced for working-age individuals than for older individuals, the overall patterns were similar (supplementary tables 3 and 4).

### Prognostic implications of late presentation

Table 3 compares the mortality outcomes from Cox models following people from the point of their first presentation over a mean 3.9 years (up to 10 years), covering 28821 deaths among 39780 individuals. Presentations were grouped as: (1) As a reference group, those who first presented with eGFR <60 but ≥45 ml/min/1.73m^2^; (2) those who previously had eGFR <60 and now newly presented with eGFR <45; and (3) those who first presented “late and unmonitored” with eGFR newly <60 and <45 occurring at the same time, excluding those with AKI. As shown in adjusted mortality curves from the fully adjusted Cox model, “late / unmonitored” presentation was associated with greater mortality than timely presentation at either <60 or <45 thresholds (figure 2). This pattern held for both deprived and affluent groups, with substantially greater mortality outcomes among those both presenting late and in deprived circumstances (supplementary figure 1). As reported in table 3, this relationship was irrespective of case mix adjustment (hazard ratio, HR 1.78, 1.70-1.87), was more pronounced over the first year of follow-up (HR 2.55, 2.36-2.75), but still evident across long term follow-up (HR 1.41, 1.32-1.51).

**Figure 2:**
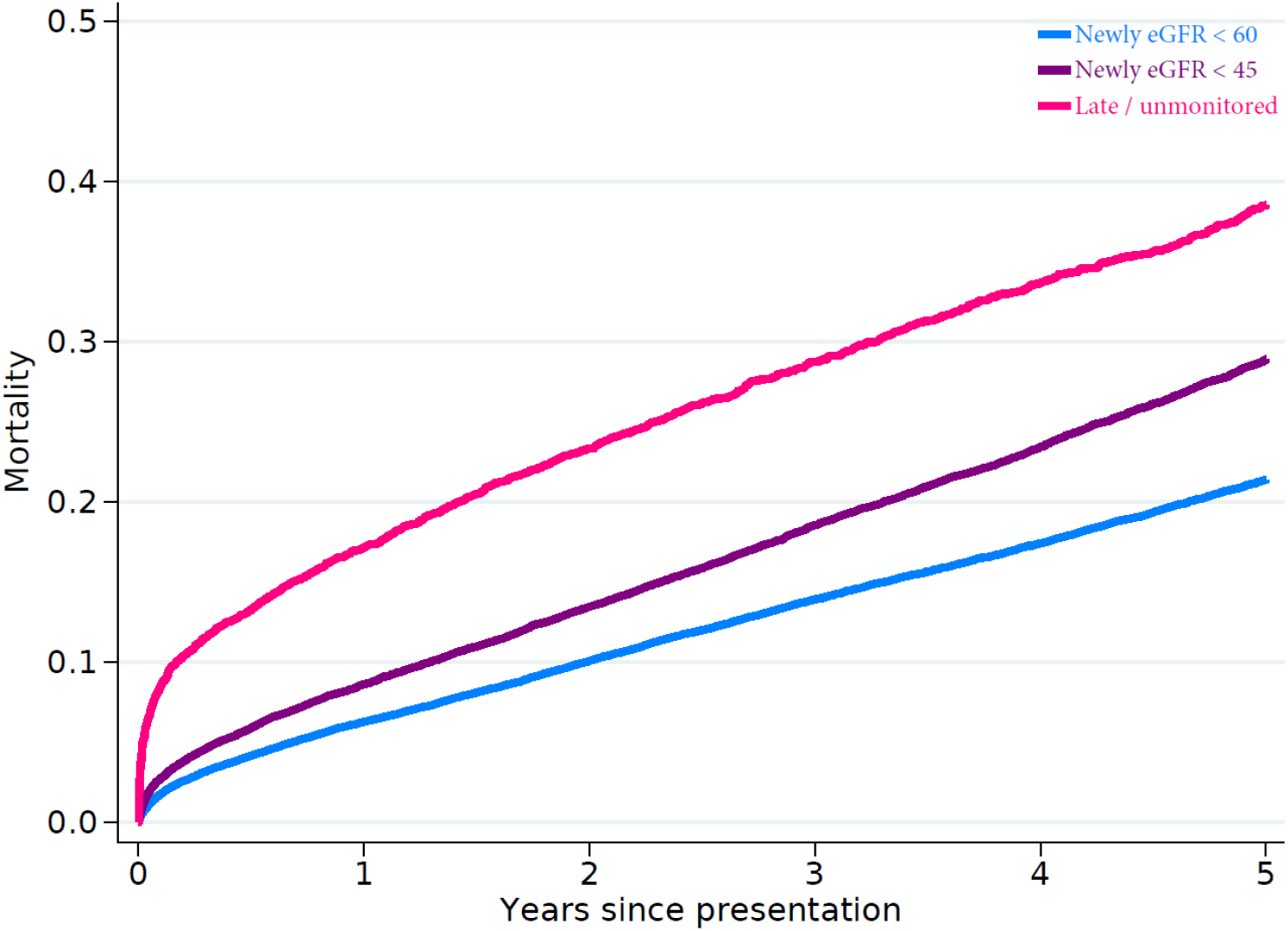
Mortality of people newly presenting at eGFR thresholds of 60 and 45, and “late / unmonitored” with < 45 ml/min/1.73m^2^

**Table 3:**
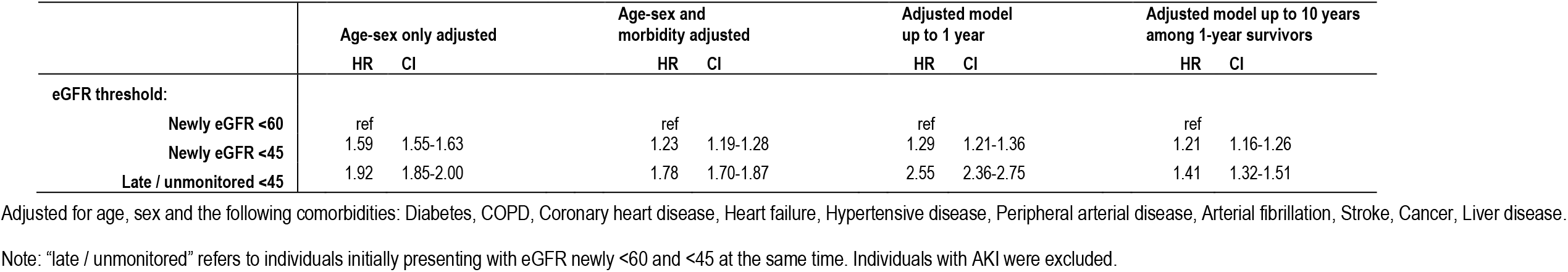
Mortality outcomes people newly presenting at eGFR thresholds of 60 and 45, and “late / unmonitored” with < 45 ml/min/1.73m. ^2^

|  | **Age-sex only adjusted** |  | **Age-sex and morbidity adjusted** |  | **Adjusted model up to 1 year** |  | **Adjusted model up to 10 years among 1-year survivors** |  |
|---|---|---|---|---|---|---|---|---|
|  | **HR** | **CI** | **HR** | **CI** | **HR** | **CI** | **HR** | **CI** |
| **eGFR threshold:** |  |  |  |  |  |  |  |  |
| **Newly eGFR <60** | ref |  | ref |  | ref |  | ref |  |
| **Newly eGFR <45** | 1.59 | 1.55-1.63 | 1.23 | 1.19-1.28 | 1.29 | 1.21-1.36 | 1.21 | 1.16-1.26 |
| **Late / unmonitored <45** | 1.92 | 1.85-2.00 | 1.78 | 1.70-1.87 | 2.55 | 2.36-2.75 | 1.41 | 1.32-1.51 |

Adjusted for age, sex and the following comorbidities: Diabetes, COPD, Coronary heart disease, Heart failure, Hypertensive disease, Peripheral arterial disease, Arterial fibrillation, Stroke, Cancer, Liver disease.
Note: “late / unmonitored” refers to individuals initially presenting with eGFR newly <60 and <45 at the same time. Individuals with AKI were excluded.

## Discussion

We found that socioeconomic inequities in early CKD care were concentrated at the point of initial clinical detection rather than distributed across subsequent early care components. Disparities were most evident in how individuals first became visible to services through late presentation, with larger differences in *unplanned* (patient-initiated) presentations than *planned* (clinician-initiated) care activities such as monitoring or prescribing. These findings indicate that a dominant source of inequity lies upstream of healthcare engagement, at or before the point of initial detection. Inequities were multilayered, affecting individuals experiencing household and neighbourhood deprivation, as well as those without transport, living alone, or reporting mental health conditions. Late presentation was also prognostically important, with excess mortality beyond the first year of follow-up.

These findings suggest that the inverse care law^32^ in CKD operates primarily through delayed and unequal detection rather than differential care after access has been established, and that delayed detection likely reflects the inherent *invisibility* of underserved communities rather than inequitable surveillance of individuals already identified as high-risk^33^.

A key implication is that a meaningful “left-shift” toward earlier intervention will require strategies that extend beyond optimisation of routine care for known patients. Approaches reliant on patient-initiated access are unlikely to reduce inequities if high-risk individuals remain undetected. Instead, proactive community-based case-finding and engagement strategies are likely to be needed. Additional barriers for underserved communities may include lack of documentation of risk such as hypertension, low awareness, limited health literacy, communication difficulties, reduced trust, financial pressures, and competing family/ household/work demands. In the UK, initiatives such as HIDDEN CKD demonstrate the feasibility of community-based outreach ^34^. Our findings support the need to develop, adapt and rigorously evaluate similar scalable interventions tailored to local community and neighbourhood contexts. The association of mental health with late presentation and emergency care further emphasises the need to integrate psychological and social support into early kidney care pathways.

A second key finding was a marked underuse of ACR surveillance among individuals with cardiovascular disease (10% vs 75% in diabetes). Although not socioeconomically patterned, this gap is likely to disproportionately affect deprived populations where cardiovascular disease is more prevalent. This gap may reflect differences in policy prioritisation and implementation between diabetes and cardiovascular disease in Scotland: with less emphasis on kidney health assessment for the former^35^ compared with an integrated diabetes care pathway^36^ and established national audit platform for the latter^37^. This represents a system-level gap in kidney surveillance beyond diabetes and a need for parity in risk assessment within CKM prevention strategies^6^.

Our findings are consistent with UK reports of low kidney health awareness, delays in nephrology referral^18^ and incomplete CKD coding in deprived neighbourhoods^20^. They extend international registry studies from the US and Australia, suggesting that inequitable kidney failure outcomes arise upstream of kidney replacement therapy care^38, 39^. Crucially, our analysis newly identifies late initial presentation as a key mechanism linking disadvantage to mortality in early CKD. This interpretation is supported by our parallel qualitative work describing a *“triple invisibility”* of early kidney disease in deprived communities: low public awareness, constrained access within pressured health-care systems, and non-specific early symptoms. Consequently, individuals in deprived communities described “early” kidney care occurring “late” for them^33^.

Strengths of our analysis include the large population-based cohort, linkage of individual and neighbourhood socioeconomic data, and systematic focus on the earliest, under-studied steps in kidney disease detection and care. Incident detection of kidney disease receives limited research attention but was possible here due to comprehensive population capture of all blood tests. This is distinct from alternative UK kidney research sources, where laboratory data availability and completeness varies when individuals move between different primary care practices and secondary care hospitals, precluding the feasible assessment of incident disease or continuity^40^. Limitations include a White European (UK) population (95%) precluding the assessment of ethnicity, and a pre-COVID-19 context. We did not cover other aspects of primary care such as consultations, or factors such as education or health literacy. Some additional “high-risk” factors for kidney disease, including imaging and genetic/family history, were also not captured. Finally, although late presentation was strongly associated with mortality, causality cannot be assumed, however this represents a high-risk group warranting prioritisation.

In summary, a dominant source of socioeconomic inequities in early kidney care is at or before initial clinical detection, reflected in late presentation. Without addressing inequities in detection and access, advances in CKD therapeutics risk preferentially benefiting those already visible to services, thereby widening disparities.

## Data Availability

Data access would require approval of NHS Scotland Public Benefit and Privacy Panel for Health and Social Care (HSC-PBPP). Information on how researchers may make requests to obtain similar datasets covering kidney health and Census data respectively can be obtained by contacting the Grampian Data Safe Haven at and the Electronic Data Research and Innovation Service (eDRIS) of Public Health Scotland at.

## Acknowledgements

We acknowledge the support of the Grampian Data Safe Haven (DaSH) facility within the Aberdeen Centre for Health Data Science and the associated financial support of the University of Aberdeen, and NHS Research Scotland (through NHS Grampian investment in DaSH). For more information, visit the DaSH website: http://www.abdn.ac.uk/iahs/facilities/grampian-data-safe-haven.php. The authors would like to acknowledge the support of the eDRIS Team (Public Health Scotland), in particular Fiona James, for their involvement in obtaining approvals, provisioning and linking data and for the use of the secure analytical platform within the National Safe Haven. This work contains statistical data from the National Records of Scotland (NRS) which is Crown Copyright. The use of NRS statistical data in this work does not imply the endorsement of NRS in relation to the interpretation or analysis of the statistical data. This work uses research datasets which may not exactly reproduce National Statistics aggregates. We are also grateful to Mr Mike Melvin and Ms Fumiko Nakamaru for their careful deliberations as public partners in the design and interpretation of this study.

## Authors’ Contributions

Study idea: AH (patient partner). Study design: EC, AH, TB, BK, MRD, SS. Supervision: SS. Statistical analysis input: EC, DN, MN, SS. Interpretation and drafting: All authors. Each author contributed important intellectual content during manuscript drafting or revision. SS is the guarantor.

---

**Supplementary table 1:** Processes of care and presenting circumstances.

|  | Prior OP creatinine among high risk |  |  |  | Prior ACR among high risk |  |  |  | Late / unmonitored presentation eGFR <45 (excl AKI) |  |  |  | Inpatient presentation (excl AKI) |  |  |  |
|---|---|---|---|---|---|---|---|---|---|---|---|---|---|---|---|---|
| n/N | 16401 / 18746 |  |  |  | 4720 / 18746 |  |  |  | 6042 / 39780 |  |  |  | 10074 / 39780 |  |  |  |
|  | % | Adj % | Adj RR | 95% CI | % | Adj % | Adj RR | 95% CI | % | Adj % | Adj RR | 95% CI | % | Adj % | Adj RR | 95% CI |
| **Age** |  |  |  |  |  |  |  |  |  |  |  |  |  |  |  |  |
| **18-65** | 87.9 | 87.5 | ref |  | 28.2 | 25.9 | ref |  | 17.6 | 17.4 | ref |  | 26.6 | 29 | ref |  |
| **>65** | 87.4 | 87.5 | 1 | 0.99-1.01 | 24.4 | 23.2 | 1.12 | 1.07-1.17 | 14.2 | 14.3 | 0.82 | 0.78-0.86 | 24.8 | 24 | 0.83 | 0.80-0.86 |
| **Sex** |  |  |  |  |  |  |  |  |  |  |  |  |  |  |  |  |
| **Male** | 89.2 | 88.7 | ref |  | 28.0 | 26 | ref |  | 16.2 | 16.1 | ref |  | 28.6 | 26.7 | ref |  |
| **Female** | 85.7 | 86.1 | 0.97 | 0.96-0.98 | 22.0 | 24.1 | 0.93 | 0.89-0.96 | 14.4 | 14.5 | 0.90 | 0.86-0.94 | 22.7 | 24.1 | 0.90 | 0.87-0.93 |
| **SIMD** |  |  |  |  |  |  |  |  |  |  |  |  |  |  |  |  |
| **(most affluent) 5** | 86.7 | 87 | ref |  | 23.8 | 25.1 | ref |  | 13.9 | 14.1 | ref |  | 23.9 | 24.5 | ref |  |
| **4** | 88.4 | 88.6 | 1.02 | 1.00-1.03 | 23.9 | 25.1 | 1 | 0.95-1.05 | 14.5 | 14.6 | 1.04 | 0.97-1.10 | 24.5 | 24.7 | 1.01 | 0.97-1.06 |
| **3** | 88.2 | 88 | 1.01 | 1.00-1.02 | 26.3 | 25.1 | 1 | 0.95-1.06 | 15.4 | 15.4 | 1.1 | 1.03-1.17 | 25.1 | 25.1 | 1.02 | 0.98-1.07 |
| **2** | 87.1 | 86.6 | 1 | 0.98-1.01 | 27.0 | 25.1 | 1 | 0.94-1.06 | 17.4 | 17.1 | 1.22 | 1.13-1.30 | 28.2 | 27.1 | 1.11 | 1.05-1.16 |
| **(most deprived) 1** | 85.2 | 84.4 | 0.97 | 0.94-0.99 | 30.3 | 26.6 | 1.06 | 0.97-1.16 | 20.4 | 19.3 | 1.37 | 1.23-1.53 | 32.0 | 30.1 | 1.23 | 1.14-1.33 |
| **NSSEC** |  |  |  |  |  |  |  |  |  |  |  |  |  |  |  |  |
| **Professional** | 87.5 | 87.5 | ref |  | 24.0 | 24.7 | ref |  | 14.0 | 14 | ref |  | 23.6 | 24.1 | ref |  |
| **Intermediate** | 87.9 | 88.1 | 1.01 | 0.99-1.02 | 24.3 | 25.1 | 1.02 | 0.96-1.08 | 14.3 | 14.5 | 1.03 | 0.97-1.11 | 24.0 | 24.5 | 1.01 | 0.97-1.06 |
| **Lower/technical** | 88.4 | 88 | 1.01 | 0.99-1.02 | 26.4 | 26.3 | 1.07 | 1.00-1.14 | 15.5 | 15.2 | 1.09 | 1.00-1.18 | 26.3 | 25.1 | 1.04 | 0.98-1.10 |
| **Semi-skilled** | 88.0 | 88 | 1.01 | 0.99-1.02 | 26.2 | 25.6 | 1.04 | 0.98-1.10 | 15.8 | 15.9 | 1.13 | 1.06-1.22 | 25.7 | 25.8 | 1.07 | 1.01-1.12 |
| **Unskilled** | 86.8 | 86.7 | 0.99 | 0.97-1.01 | 26.4 | 25 | 1.01 | 0.96-1.07 | 16.5 | 16.3 | 1.17 | 1.09-1.25 | 27.6 | 26.7 | 1.11 | 1.05-1.16 |
| **Unemployed** | 80.7 | 81.6 | 0.93 | 0.89-0.98 | 22.6 | 22.7 | 0.92 | 0.78-1.08 | 23.7 | 23.7 | 1.69 | 1.48-1.93 | 34.7 | 35.7 | 1.48 | 1.34-1.63 |
| **Car, Rurality** |  |  |  |  |  |  |  |  |  |  |  |  |  |  |  |  |
| **Urban, car** | 88.9 | 88.6 | ref |  | 26.2 | 25.8 | ref |  | 14.2 | 14 | ref |  | 24.2 | 24 | ref |  |
| **Urban, no car** | 82.9 | 83.5 | 0.94 | 0.93-0.96 | 24.0 | 24.7 | 0.96 | 0.91-1.01 | 19.5 | 20.1 | 1.43 | 1.35-1.52 | 31.9 | 31.6 | 1.32 | 1.27-1.37 |
| **Rural, car** | 89.2 | 89.1 | 1.01 | 0.99-1.02 | 24.7 | 24.8 | 0.96 | 0.92-1.00 | 13.6 | 13.5 | 0.96 | 0.91-1.02 | 22.3 | 22.8 | 0.95 | 0.92-0.99 |
| **Rural, no car** | 81.6 | 82.6 | 0.93 | 0.90-0.97 | 21.1 | 22 | 0.85 | 0.76-0.96 | 18.9 | 20.5 | 1.46 | 1.30-1.63 | 28.9 | 30.1 | 1.26 | 1.15-1.37 |
| **Other Context** |  |  |  |  |  |  |  |  |  |  |  |  |  |  |  |  |
| **Doesn’t live alone** | 88.8 | 88.4 | ref |  | 26.2 | 25.4 | ref |  | 14.2 | 13.9 | ref |  | 24.0 | 23.7 | ref |  |
| **Lives alone** | 84.4 | 85.5 | 0.96 | 0.95-0.98 | 22.7 | 24.5 | 0.96 | 0.92-1.01 | 17.7 | 18.6 | 1.34 | 1.27-1.41 | 28.8 | 29.8 | 1.26 | 1.21-1.30 |
| **No mental health** | 87.5 | 87.6 | ref |  | 25.0 | 25.3 | ref |  | 15.0 | 15 | ref |  | 25.0 | 25.1 | ref |  |
| **Mental health** | 86.9 | 86 | 0.98 | 0.96-1.01 | 28.6 | 23.3 | 0.92 | 0.84-1.01 | 19.8 | 18.3 | 1.22 | 1.10-1.35 | 32.0 | 30.5 | 1.21 | 1.13-1.31 |
| **No diabetes** | 85.1 | 85.1 | ref |  | 9.6 | 9.6 | ref |  | 14.8 | 14.9 | ref |  | 24.1 | 24.8 | ref |  |
| **Diabetes** | 95.3 | 95.1 | 1.12 | 1.11-1.13 | 75.3 | 75.3 | 7.84 | 7.43-8.27 | 19.0 | 18 | 1.21 | 1.12-1.30 | 37.5 | 28.7 | 1.16 | 1.10-1.21 |

Adjusted % and RRs are derived from modified Poisson GLM models with robust standard errors; adjusted for age, sex and the following comorbidities: Diabetes, COPD, Coronary heart disease, Heart failure, Hypertensive disease, Peripheral arterial disease, Arterial fibrillation, Stroke, Cancer, Liver disease (except for where the respective variable is to be left out)
High risk = History of diabetes or vascular risk factor that warrants regular screening

**Supplementary table 2:** Processes of care and subsequent care after presentation.

|  | Repeat test 14d |  |  |  | Test for chronicity after 90d |  |  |  | Available ACR test |  |  |  | RAASi among indicated |  |  |  | ED attendance |  |  |  |
|---|---|---|---|---|---|---|---|---|---|---|---|---|---|---|---|---|---|---|---|---|
| n/N | 9375 / 29823 |  |  |  | 26467 / 38462 |  |  |  | 9755 / 38462 |  |  |  | 1057 / 1523 |  |  |  | 4131 / 25084 |  |  |  |
|  | % | Adj % | Adj RR | 95% CI | % | Adj % | Adj RR | 95% CI | % | Adj % | Adj RR | 95% CI | % | Adj % | Adj RR | 95% CI | % | Adj % | Adj RR | 95% CI |
| **Age** |  |  |  |  |  |  |  |  |  |  |  |  |  |  |  |  |  |  |  |  |
| **18-65** | 39.3 | 39.6 | ref |  | 66.5 | 67.7 | ref |  | 23.1 | 22.9 | ref |  | 69.8 | 72.1 | ref |  | 16.2 | 17.1 | ref |  |
| **>65** | 28.3 | 28.2 | 0.71 | 0.69-0.74 | 69.9 | 69.3 | 1.02 | 1.01-1.04 | 26.4 | 26.5 | 1.15 | 1.11-1.20 | 69.2 | 68.2 | 0.95 | 0.88-1.02 | 16.6 | 16.2 | 0.94 | 0.89-1.00 |
| **Sex** |  |  |  |  |  |  |  |  |  |  |  |  |  |  |  |  |  |  |  |  |
| **Male** | 33.8 | 33.7 | ref |  | 71.5 | 70.2 | ref |  | 28.8 | 26.9 | ref |  | 71.7 | 71.5 | ref |  | 16.6 | 15.9 | ref |  |
| **Female** | 29.8 | 29.8 | 0.89 | 0.86-0.92 | 66.7 | 67.7 | 0.96 | 0.95-0.98 | 22.6 | 24 | 0.89 | 0.86-0.92 | 65.0 | 65.3 | 0.91 | 0.92-0.98 | 16.3 | 17 | 1.07 | 1.01-1.13 |
| **SIMD** |  |  |  |  |  |  |  |  |  |  |  |  |  |  |  |  |  |  |  |  |
| **(most affluent) 5** | 31.1 | 31.5 | ref |  | 67.8 | 68.1 | ref |  | 23.7 | 24.3 | ref |  | 67.1 | 67.3 | ref |  | 15.7 | 16.1 | ref |  |
| **4** | 29.8 | 29.9 | 0.95 | 0.91-0.99 | 68.7 | 68.9 | 1.01 | 1.00-1.03 | 24.5 | 25 | 1.03 | 0.99-1.07 | 70.8 | 71 | 1.06 | 0.97-1.15 | 15.6 | 15.8 | 0.98 | 0.91-1.06 |
| **3** | 31.2 | 31.3 | 0.99 | 0.95-1.04 | 69.5 | 69.4 | 1.02 | 1.00-1.04 | 26.4 | 25.8 | 1.06 | 1.01-1.11 | 67.6 | 67.3 | 1 | 0.91-1.10 | 16.3 | 16.1 | 1 | 0.93-1.09 |
| **2** | 34.2 | 33.5 | 1.06 | 1.01-1.12 | 69.6 | 69.1 | 1.02 | 0.99-1.04 | 27.8 | 26.7 | 1.1 | 1.05-1.15 | 74.4 | 74.1 | 1.1 | 1.00-1.21 | 18.6 | 17.9 | 1.11 | 1.02-1.21 |
| **(most deprived) 1** | 37.6 | 35.1 | 1.12 | 1.03-1.21 | 70.0 | 69.4 | 1.02 | 0.98-1.06 | 29.2 | 27.5 | 1.13 | 1.05-1.22 | 65.3 | 65.7 | 0.98 | 0.83-1.15 | 21.2 | 20.2 | 1.26 | 1.11-1.44 |
| **NSSEC** |  |  |  |  |  |  |  |  |  |  |  |  |  |  |  |  |  |  |  |  |
| **Professional** | 31.5 | 30.8 | ref |  | 67.8 | 68 | ref |  | 24.5 | 25.1 | ref |  | 71.2 | 70.7 | ref |  | 15.4 | 15.7 | ref |  |
| **Intermediate** | 30.1 | 30.7 | 1 | 0.95-1.04 | 67.8 | 68 | 1 | 0.98-1.02 | 24.7 | 25.1 | 1 | 0.96-1.04 | 68.0 | 67.1 | 0.95 | 0.86-1.05 | 15.2 | 15.4 | 0.98 | 0.90-1.06 |
| **Lower/technical** | 33.4 | 32.5 | 1.05 | 1.00-1.11 | 69.8 | 69.4 | 1.02 | 0.97-1.04 | 25.3 | 25.1 | 1 | 0.95-1.05 | 72.1 | 71.5 | 1.01 | 0.91-1.12 | 17.2 | 17.1 | 1.09 | 0.99-1.20 |
| **Semi-skilled** | 30.9 | 31.6 | 1.03 | 0.98-1.08 | 70.0 | 70 | 1.03 | 1.01-1.05 | 25.8 | 25.5 | 1.01 | 0.97-1.07 | 66.5 | 66.6 | 0.94 | 0.85-1.05 | 17.6 | 17.2 | 1.1 | 1.01-1.20 |
| **Unskilled** | 32.6 | 32.8 | 1.07 | 1.02-1.12 | 70.2 | 69.7 | 1.02 | 1.00-1.04 | 27.3 | 26.1 | 1.04 | 0.99-1.09 | 69.7 | 71.2 | 1.01 | 0.92-1.10 | 17.6 | 17.1 | 1.09 | 1.00-1.18 |
| **Unemployed** | 27.2 | 28.5 | 0.92 | 0.80-1.06 | 67.2 | 68.2 | 1 | 0.95-1.05 | 23.6 | 24.1 | 0.96 | 0.85-1.08 | 62.1 | 66 | 0.93 | 0.69-1.26 | 23.7 | 23.5 | 1.5 | 1.26-1.78 |
| **Car, Rurality** |  |  |  |  |  |  |  |  |  |  |  |  |  |  |  |  |  |  |  |  |
| **Urban, car** | 32.6 | 31.8 | ref |  | 69.2 | 69 | ref |  | 25.8 | 25.6 | ref |  | 70.7 | 70 | ref |  | 16.4 | 16.4 | ref |  |
| **Urban, no car** | 31.3 | 34.1 | 1.07 | 1.02-1.12 | 67.2 | 67.2 | 0.97 | 0.96-0.99 | 24.6 | 24.6 | 0.96 | 0.82-1.01 | 64.2 | 65.6 | 0.94 | 0.85-1.03 | 21.7 | 21 | 1.28 | 1.19-1.38 |
| **Rural, car** | 30.0 | 29.3 | 0.92 | 0.89-0.96 | 69.1 | 69.4 | 1.01 | 0.99-1.02 | 25.3 | 25.6 | 1 | 0.97-1.04 | 72.8 | 72.6 | 1.04 | 0.96-1.12 | 13.4 | 13.7 | 0.83 | 0.78-0.90 |
| **Rural, no car** | 26.5 | 30.8 | 0.97 | 0.87-1.07 | 69.6 | 70 | 1.01 | 0.98-1.06 | 23.4 | 23.6 | 0.92 | 0.84-1.01 | 50.0 | 54.1 | 0.77 | 0.57-1.05 | 20.4 | 20 | 1.22 | 1.05-1.43 |
| **Other Context** |  |  |  |  |  |  |  |  |  |  |  |  |  |  |  |  |  |  |  |  |
| **Doesn’t live alone** | 32.5 | 31.3 | ref |  | 69.0 | 68.9 | ref |  | 25.9 | 25.7 | ref |  | 72.6 | 72 | ref |  | 15.9 | 15.9 | ref |  |
| **Lives alone** | 28.4 | 31.7 | 1 | 0.96-1.05 | 68.2 | 68.5 | 0.99 | 0.98-1.01 | 23.9 | 24.3 | 0.95 | 0.91-0.98 | 59.9 | 62 | 0.86 | 0.78-0.94 | 18.4 | 18.3 | 1.16 | 1.09-1.23 |
| **No mental health** | 31.3 | 31.5 | ref |  | 68.7 | 68.7 | ref |  | 25.4 | 25.4 | ref |  | 70.2 | 70.1 | ref |  | 16.1 | 16.1 | ref |  |
| **Mental health** | 34.6 | 30.8 | 0.98 | 0.90-1.06 | 71.7 | 72.3 | 1.05 | 1.02-1.09 | 23.6 | 22.7 | 0.89 | 0.82-0.96 | 54.2 | 55 | 0.78 | 0.63-0.97 | 25.6 | 24.7 | 1.54 | 1.38-1.72 |
| **No diabetes** | 30.9 | 31.1 | ref |  | 67.0 | 67.5 | ref |  | 19.2 | 19.1 | ref |  | 70.1 | 71.8 | ref |  | 15.6 | 16 | ref |  |
| **Diabetes** | 37.4 | 35.4 | 1.14 | 1.07-1.21 | 86.2 | 80.6 | 1.19 | 1.18-1.21 | 84.9 | 86.8 | 4.54 | 4.41-4.68 | 69.3 | 69 | 0.96 | 0.87-1.06 | 23.8 | 20 | 1.26 | 1.16-1.36 |

Adjusted % and RRs are derived from modified Poisson GLM models with robust standard errors; adjusted for age, sex and the following comorbidities: Diabetes, COPD, Coronary heart disease, Heart failure, Hypertensive disease, Peripheral arterial disease, Arterial fibrillation, Stroke, Cancer, Liver disease (except for where the respective variable is to be left out)
RAASi indication = diabetes & urine ACR >3; or hypertension & urine ACR >30; or urine ACR >70

**Supplementary table 3:**
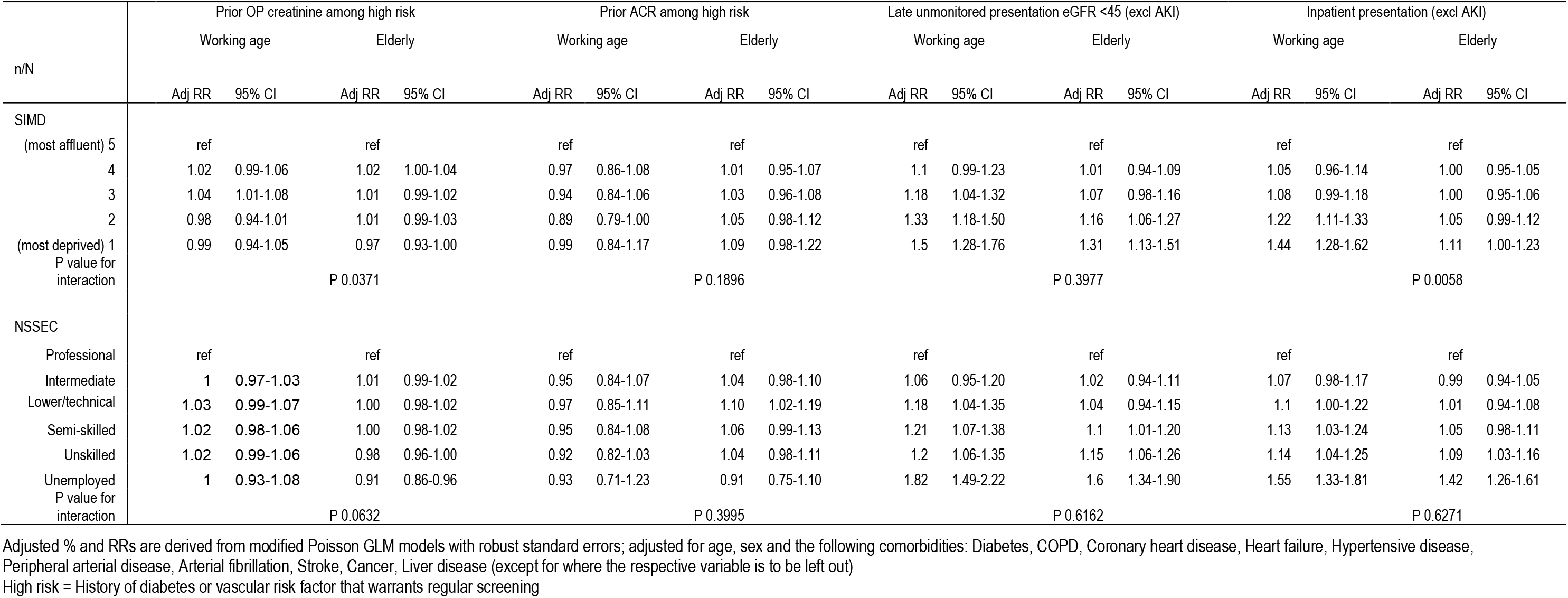
Presenting circumstances stratified by age.

| n/N | Prior OP creatinine among high risk |  |  |  | Prior ACR among high risk |  |  |  | Late unmonitored presentation eGFR <45 (excl AKI) |  |  |  | Inpatient presentation (excl AKI) |  |  |  |
|---|---|---|---|---|---|---|---|---|---|---|---|---|---|---|---|---|
|  | Working age |  | Elderly |  | Working age |  | Elderly |  | Working age |  | Elderly |  | Working age |  | Elderly |  |
|  | Adj RR | 95% CI | Adj RR | 95% CI | Adj RR | 95% CI | Adj RR | 95% CI | Adj RR | 95% CI | Adj RR | 95% CI | Adj RR | 95% CI | Adj RR | 95% CI |
| SIMD |  |  |  |  |  |  |  |  |  |  |  |  |  |  |  |  |
| (most affluent) 5 | ref |  | ref |  | ref |  | ref |  | ref |  | ref |  | ref |  | ref |  |
| 4 | 1.02 | 0.99-1.06 | 1.02 | 1.00-1.04 | 0.97 | 0.86-1.08 | 1.01 | 0.95-1.07 | 1.1 | 0.99-1.23 | 1.01 | 0.94-1.09 | 1.05 | 0.96-1.14 | 1.00 | 0.95-1.05 |
| 3 | 1.04 | 1.01-1.08 | 1.01 | 0.99-1.02 | 0.94 | 0.84-1.06 | 1.03 | 0.96-1.08 | 1.18 | 1.04-1.32 | 1.07 | 0.98-1.16 | 1.08 | 0.99-1.18 | 1.00 | 0.95-1.06 |
| 2 | 0.98 | 0.94-1.01 | 1.01 | 0.99-1.03 | 0.89 | 0.79-1.00 | 1.05 | 0.98-1.12 | 1.33 | 1.18-1.50 | 1.16 | 1.06-1.27 | 1.22 | 1.11-1.33 | 1.05 | 0.99-1.12 |
| (most deprived) 1 | 0.99 | 0.94-1.05 | 0.97 | 0.93-1.00 | 0.99 | 0.84-1.17 | 1.09 | 0.98-1.22 | 1.5 | 1.28-1.76 | 1.31 | 1.13-1.51 | 1.44 | 1.28-1.62 | 1.11 | 1.00-1.23 |
| P value for interaction |  |  | P 0.0371 |  |  |  | P 0.1896 |  |  |  | P 0.3977 |  |  |  | P 0.0058 |  |
| NSSEC |  |  |  |  |  |  |  |  |  |  |  |  |  |  |  |  |
| Professional | ref |  | ref |  | ref |  | ref |  | ref |  | ref |  | ref |  | ref |  |
| Intermediate | 1 | 0.97-1.03 | 1.01 | 0.99-1.02 | 0.95 | 0.84-1.07 | 1.04 | 0.98-1.10 | 1.06 | 0.95-1.20 | 1.02 | 0.94-1.11 | 1.07 | 0.98-1.17 | 0.99 | 0.94-1.05 |
| Lower/technical | 1.03 | 0.99-1.07 | 1.00 | 0.98-1.02 | 0.97 | 0.85-1.11 | 1.10 | 1.02-1.19 | 1.18 | 1.04-1.35 | 1.04 | 0.94-1.15 | 1.1 | 1.00-1.22 | 1.01 | 0.94-1.08 |
| Semi-skilled | 1.02 | 0.98-1.06 | 1.00 | 0.98-1.02 | 0.95 | 0.84-1.08 | 1.06 | 0.99-1.13 | 1.21 | 1.07-1.38 | 1.1 | 1.01-1.20 | 1.13 | 1.03-1.24 | 1.05 | 0.98-1.11 |
| Unskilled | 1.02 | 0.99-1.06 | 0.98 | 0.96-1.00 | 0.92 | 0.82-1.03 | 1.04 | 0.98-1.11 | 1.2 | 1.06-1.35 | 1.15 | 1.06-1.26 | 1.14 | 1.04-1.25 | 1.09 | 1.03-1.16 |
| Unemployed | 1 | 0.93-1.08 | 0.91 | 0.86-0.96 | 0.93 | 0.71-1.23 | 0.91 | 0.75-1.10 | 1.82 | 1.49-2.22 | 1.6 | 1.34-1.90 | 1.55 | 1.33-1.81 | 1.42 | 1.26-1.61 |
| P value for interaction |  |  | P 0.0632 |  |  |  | P 0.3995 |  |  |  | P 0.6162 |  |  |  | P 0.6271 |  |

Adjusted % and RRs are derived from modified Poisson GLM models with robust standard errors; adjusted for age, sex and the following comorbidities: Diabetes, COPD, Coronary heart disease, Heart failure, Hypertensive disease, Peripheral arterial disease, Arterial fibrillation, Stroke, Cancer, Liver disease (except for where the respective variable is to be left out)
High risk = History of diabetes or vascular risk factor that warrants regular screening

**Supplementary table 4:**
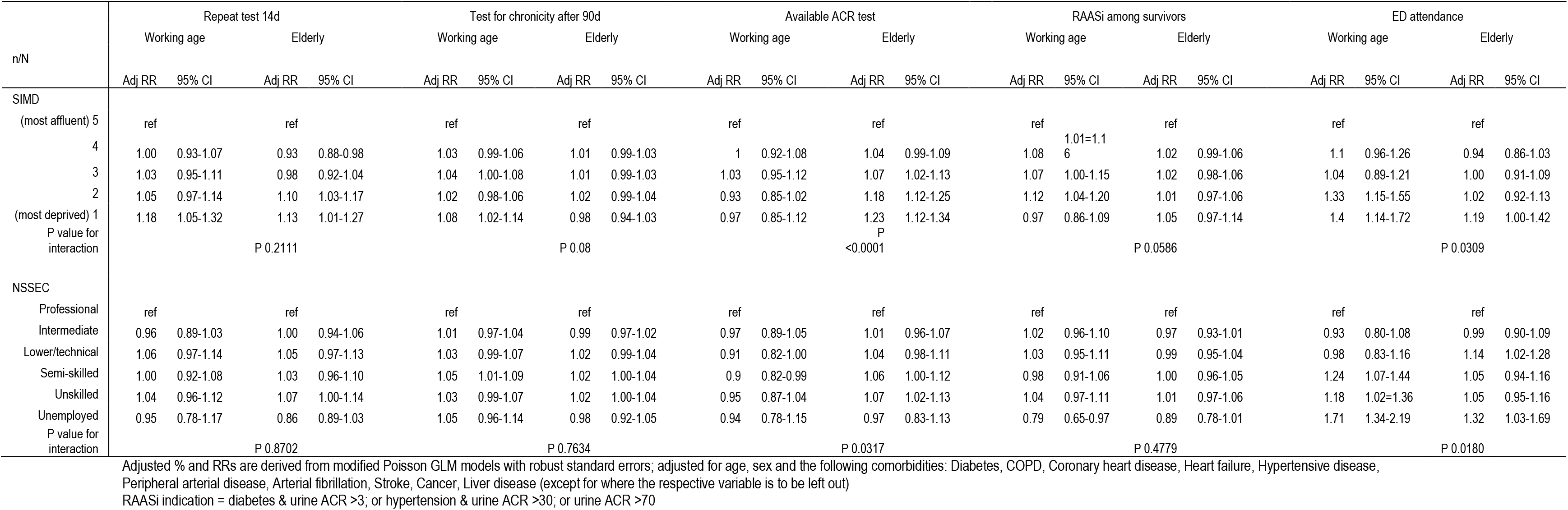
Subsequent care stratified by age.

|  | Repeat test 14d |  |  |  | Test for chronicity after 90d |  |  |  | Available ACR test |  |  |  | RAASi among survivors |  |  |  | ED attendance |  |  |  |
|---|---|---|---|---|---|---|---|---|---|---|---|---|---|---|---|---|---|---|---|---|
| n/N | Working age |  | Elderly |  | Working age |  | Elderly |  | Working age |  | Elderly |  | Working age |  | Elderly |  | Working age |  | Elderly |  |
|  | Adj RR | 95% CI | Adj RR | 95% CI | Adj RR | 95% CI | Adj RR | 95% CI | Adj RR | 95% CI | Adj RR | 95% CI | Adj RR | 95% CI | Adj RR | 95% CI | Adj RR | 95% CI | Adj RR | 95% CI |
| SIMD |  |  |  |  |  |  |  |  |  |  |  |  |  |  |  |  |  |  |  |  |
| (most affluent) 5 | ref |  | ref |  | ref |  | ref |  | ref |  | ref |  | ref |  | ref |  | ref |  | ref |  |
| 4 | 1.00 | 0.93-1.07 | 0.93 | 0.88-0.98 | 1.03 | 0.99-1.06 | 1.01 | 0.99-1.03 | 1 | 0.92-1.08 | 1.04 | 0.99-1.09 | 1.08 | 1.01=1.16 | 1.02 | 0.99-1.06 | 1.1 | 0.96-1.26 | 0.94 | 0.86-1.03 |
| 3 | 1.03 | 0.95-1.11 | 0.98 | 0.92-1.04 | 1.04 | 1.00-1.08 | 1.01 | 0.99-1.03 | 1.03 | 0.95-1.12 | 1.07 | 1.02-1.13 | 1.07 | 1.00-1.15 | 1.02 | 0.98-1.06 | 1.04 | 0.89-1.21 | 1.00 | 0.91-1.09 |
| 2 | 1.05 | 0.97-1.14 | 1.10 | 1.03-1.17 | 1.02 | 0.98-1.06 | 1.02 | 0.99-1.04 | 0.93 | 0.85-1.02 | 1.18 | 1.12-1.25 | 1.12 | 1.04-1.20 | 1.01 | 0.97-1.06 | 1.33 | 1.15-1.55 | 1.02 | 0.92-1.13 |
| (most deprived) 1 | 1.18 | 1.05-1.32 | 1.13 | 1.01-1.27 | 1.08 | 1.02-1.14 | 0.98 | 0.94-1.03 | 0.97 | 0.85-1.12 | 1.23 | 1.12-1.34 | 0.97 | 0.86-1.09 | 1.05 | 0.97-1.14 | 1.4 | 1.14-1.72 | 1.19 | 1.00-1.42 |
| P value for interaction |  |  | P 0.2111 |  |  |  | P 0.08 |  |  |  | P <0.0001 |  |  |  | P 0.0586 |  |  |  | P 0.0309 |  |
| NSSEC |  |  |  |  |  |  |  |  |  |  |  |  |  |  |  |  |  |  |  |  |
| Professional | ref |  | ref |  | ref |  | ref |  | ref |  | ref |  | ref |  | ref |  | ref |  | ref |  |
| Intermediate | 0.96 | 0.89-1.03 | 1.00 | 0.94-1.06 | 1.01 | 0.97-1.04 | 0.99 | 0.97-1.02 | 0.97 | 0.89-1.05 | 1.01 | 0.96-1.07 | 1.02 | 0.96-1.10 | 0.97 | 0.93-1.01 | 0.93 | 0.80-1.08 | 0.99 | 0.90-1.09 |
| Lower/technical | 1.06 | 0.97-1.14 | 1.05 | 0.97-1.13 | 1.03 | 0.99-1.07 | 1.02 | 0.99-1.04 | 0.91 | 0.82-1.00 | 1.04 | 0.98-1.11 | 1.03 | 0.95-1.11 | 0.99 | 0.95-1.04 | 0.98 | 0.83-1.16 | 1.14 | 1.02-1.28 |
| Semi-skilled | 1.00 | 0.92-1.08 | 1.03 | 0.96-1.10 | 1.05 | 1.01-1.09 | 1.02 | 1.00-1.04 | 0.9 | 0.82-0.99 | 1.06 | 1.00-1.12 | 0.98 | 0.91-1.06 | 1.00 | 0.96-1.05 | 1.24 | 1.07-1.44 | 1.05 | 0.94-1.16 |
| Unskilled | 1.04 | 0.96-1.12 | 1.07 | 1.00-1.14 | 1.03 | 0.99-1.07 | 1.02 | 1.00-1.04 | 0.95 | 0.87-1.04 | 1.07 | 1.02-1.13 | 1.04 | 0.97-1.11 | 1.01 | 0.97-1.06 | 1.18 | 1.02=1.36 | 1.05 | 0.95-1.16 |
| Unemployed | 0.95 | 0.78-1.17 | 0.86 | 0.89-1.03 | 1.05 | 0.96-1.14 | 0.98 | 0.92-1.05 | 0.94 | 0.78-1.15 | 0.97 | 0.83-1.13 | 0.79 | 0.65-0.97 | 0.89 | 0.78-1.01 | 1.71 | 1.34-2.19 | 1.32 | 1.03-1.69 |
| P value for interaction |  |  | P 0.8702 |  |  |  | P 0.7634 |  |  |  | P 0.0317 |  |  |  | P 0.4779 |  |  |  | P 0.0180 |  |

Adjusted % and RRs are derived from modified Poisson GLM models with robust standard errors; adjusted for age, sex and the following comorbidities: Diabetes, COPD, Coronary heart disease, Heart failure, Hypertensive disease, Peripheral arterial disease, Arterial fibrillation, Stroke, Cancer, Liver disease (except for where the respective variable is to be left out)
RAASi indication = diabetes & urine ACR >3; or hypertension & urine ACR >30; or urine ACR >70

**Supplementary figure 1:**
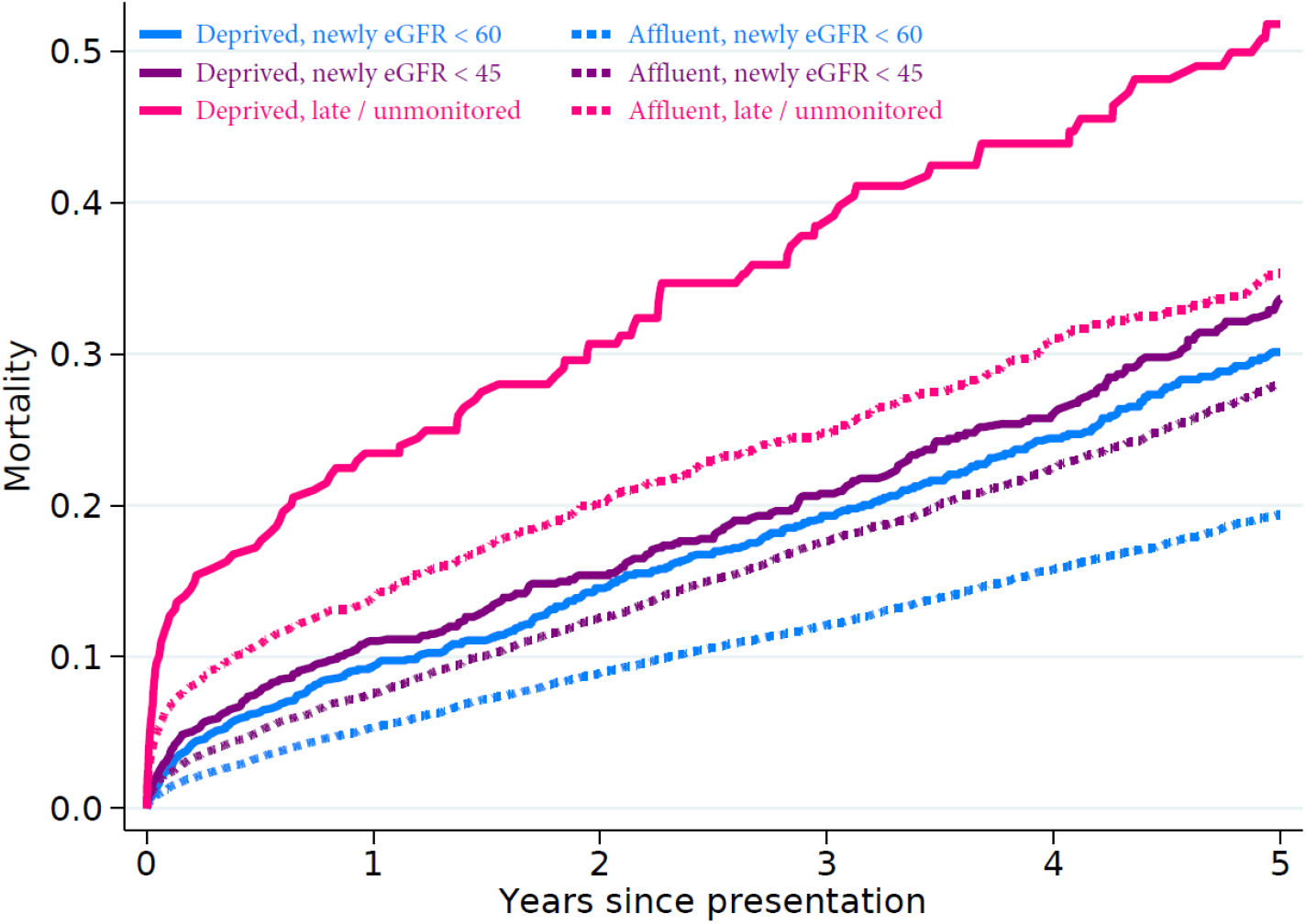
Mortality of people newly presenting at eGFR thresholds, within deprived and affluent neighbourhoods

